# Synthesizing Contrast-Enhanced T1 MR Image Using Multiparametric Sequences and Attention to Brain Tumor

**DOI:** 10.1101/2025.11.14.25340220

**Authors:** Ziheng Guo, Boyang Pan, Aijing Lin, Nan-Jie Gong

## Abstract

**Objective:** The administration of gadolinium-based contrast agents (GBCAs) for acquiring contrast-enhanced T1-weighted magnetic resonance imaging (T1C MRI) is associated with potential safety risks, including tissue deposition and nephrogenic systemic fibrosis. This work aims to develop a deep learning framework for synthesizing high-fidelity T1C MRI directly from multi-parametric, non-contrast sequences, thereby eliminating the need for GBCAs while preserving critical diagnostic information for brain tumor assessment.

**Methods:** We propose ZeroCEMR, a novel two-stage deep learning framework for GBCA-free T1C MRI synthesis. The model was developed and evaluated using multi-institutional datasets, including the BraTS 2021 benchmark and a clinical cohort. The first stage employs a Global Anatomical Encoder and YOLO-based tumor detectors to extract whole-brain structural features from T1-weighted (T1w), T2-weighted (T2w), and Fluid-Attenuated Inversion Recovery (FLAIR) images. These global and tumor-specific features are fused and decoded to generate a preliminary T1C image. The second stage refines this initial prediction by incorporating Diffusion-Weighted Imaging (DWI) and Apparent Diffusion Coefficient (ADC) sequences. Key innovations include an inter-stage residual fusion mechanism, a Squeeze-and-Excitation attention block for enhanced lesion sensitivity, and a multi-scale Feature Pyramid Network integrated within a U-Net architecture to accurately capture both macroscopic anatomy and microscopic pathological details.

**Results:** The proposed ZeroCEMR framework achieved superior performance, with a peak signal-to-noise ratio (PSNR) of 37.82 and a structural similarity index measure (SSIM) of 0.98, significantly outperforming existing methods. Ablation studies confirmed the individual contribution of each architectural component, demonstrating progressive improvements in both quantitative metrics and qualitative lesion delineation.

**Conclusion:** ZeroCEMR establishes a new state-of-the-art framework for synthetic T1C MRI generation. By effectively leveraging multi-parametric data and a lesion-focused architecture, our framework generates clinically viable contrast-enhanced images without GBCA administration. This approach represents a significant step towards safer, quantitative neuro-oncological imaging workflows.

## Introduction

Contrast-enhanced MRI (CEMRI) is critical for the detection, characterization, and monitoring of brain tumors, as the administration of gadolinium-based contrast agents (GBCA) highlights regions of blood-brain barrier disruption associated with pathological tissue [1, 2]. However, the use of GBCAs pose significant clinical risks. These include the potential for Nephrogenic Systemic Fibrosis (NSF) in patients with renal impairment [3] and the long-term deposition of gadolinium in body tissues, including the brain, with uncertain consequences [4, 5]. These safety concerns have prompted regulatory agencies, such as the European Medicines Agency (EMA) to restrict the use of certain linear gadolinium agents in 2017 [6]. Furthermore, the injection of contrast agents increases examination costs, procedural complexity, and patient discomfort, limiting its applicability in specific populations, such as those with allergies or requiring repeated scans.

To address these issues, synthesizing CEMRI from non-contrast multi-modality MRI data has emerged as a promising research direction [7,8,9]. This approach aims to computationally generate synthetic CEMRI that replicate the diagnostic information of acquired contrast-enhanced scans, thereby eliminating the need for GBCA administration. Early deep learning-based methods have demonstrated the feasibility of this task, primarily utilizing Convolutional Neural Networks (CNNs) [10–13] and Generative Adversarial Networks (GANs) [14] to learn the mapping from pre-contrast sequences like T1-weighted (T1w), T2-weighted (T2w), and Fluid-Attenuated Inversion Recovery (FLAIR) to their post-contrast counterparts.

Despite these advances, existing methods exhibit critical shortcomings, particularly in achieving high sensitivity for tumor detection. Synthesized images often suffer from blurred pathological representations and inadequate delineation of small or complex lesions[8]. This lack of sensitivity can be attributed to several factors: (1) the dilution of subtle tumor features within the global anatomical context when processed by standard encoder-decoder networks; (2) insufficient utilization of complementary diagnostic information, especially from diffusion-weighted sequences; and (3) an inability to effectively focus the synthesis process on clinically critical regions.

Aiming to address these challenges, we introduce ZeroCEMR, a novel two-stage framework for synthesizing high-quality CEMRI from multi-parametric MRI data without contrast agents. The key innovations of this framework are:

1. Integration of DWI and ADC data: We leverage a comprehensive multi-parametric MRI protocol, incorporating five distinct non-contrast sequences: T1w, T2w, FLAIR, Diffusion-Weighted Imaging (DWI), and Apparent Diffusion Coefficient (ADC). The T1w, T2w, and FLAIR sequences provide high-resolution anatomical and structural context, which is essential for generating the overall architectural integrity of the synthetic CEMRI. Concurrently, the DWI and ADC sequences contribute critical diffusion-based biomarkers that characterize tissue cellularity and microstructural integrity, thereby enhancing the fidelity of contrast representation and improving the delineation of pathological regions in the synthesized images.
2. YOLO-based tumor detection: We introduce a dedicated Multi-Modal YOLO-based tumor detection module that explicitly localizes lesion regions across different input sequences. This ensures that tumor-specific features are preserved and enhanced, preventing their dilution during global feature extraction.
3. Two-stage architecture: Our framework employs a coarse-to-fine generation strategy. The first stage produces an initial T1C prediction from T1w, T2w, and FLAIR images, while the second stage refines this prediction by incorporating DWI and ADC data, utilizing a Feature Pyramid Network (FPN) and attention mechanisms to improve boundary precision and lesion conspicuity.

In this work, we investigated the practicability of our two-stage framework, tested its performance and compared it to existing model available.

## 2. Methodology

In this section, we first delineate the overall architecture of ZeroCEMR framework. Then we describe our key architectural components: Multi-Modal YOLO Tumor Detection Module, Inter-Stage Residual Fusion, Tumor Feature Attention Module (SE-Block) and Multi-Scale Feature Pyramid Integrated UNet. Finally, we detail the loss functions.

### 2.1. Overview of the ZeroCEMR Framework

As illustrated in Fig. 1., the overall ZeroCEMR framework employs a two-stage cascaded architecture to integrate multi-modal MRI data and deep neural networks for T1 contrast-enhanced (T1C) image generation without contrast agents.

**Fig. 1.**
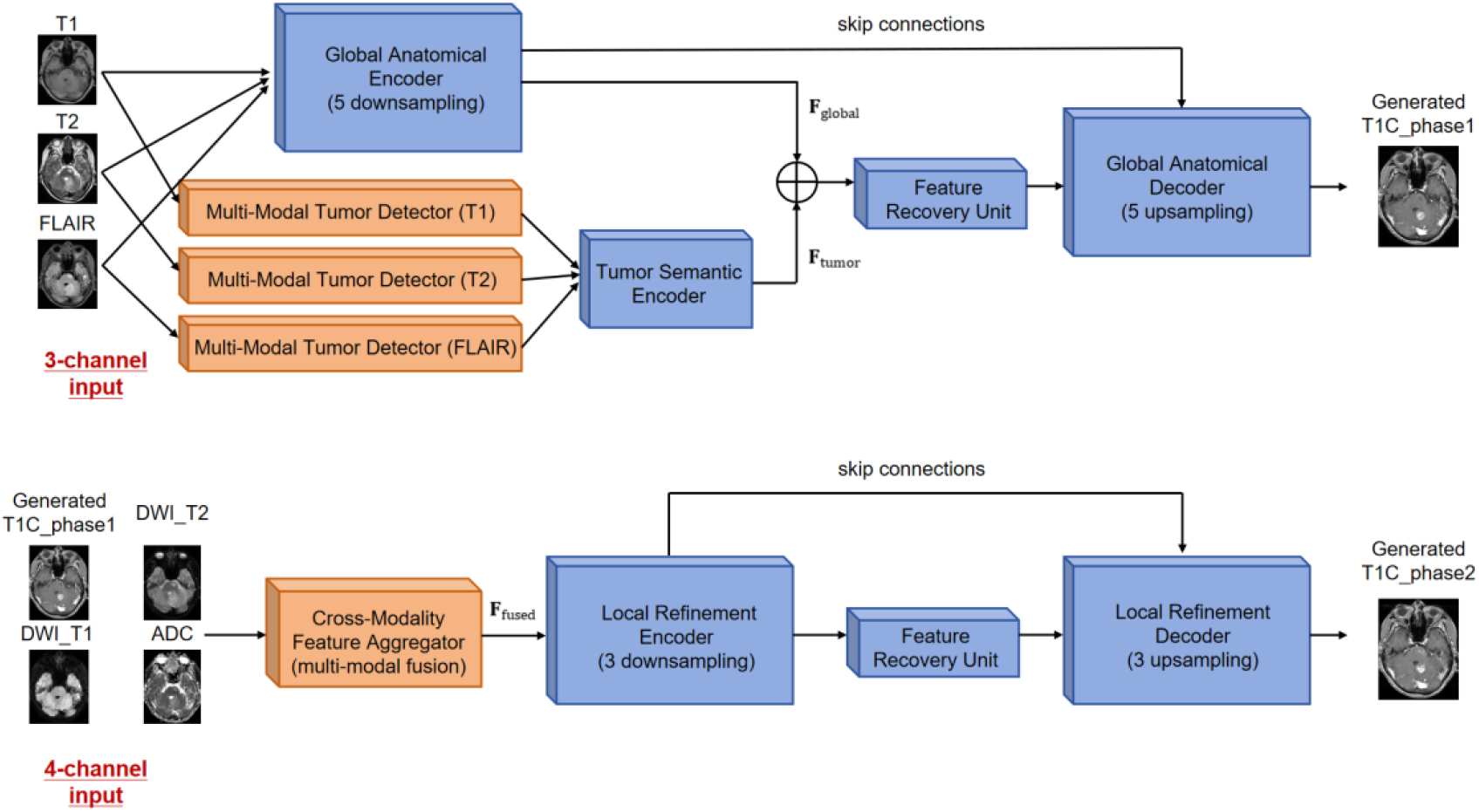
Two-stage architecture of the ZeroCEMR framework

#### Stage 1: Multi-Modal Feature Extraction and Preliminary Generation

The input modalities for stage 1 comprise T1w, T2w, and FLAIR images, each with a spatial resolution of 512×512. At this stage, a modality-specific detection-encoding pipeline was implemented to simultaneously capture global brain structures and tumor-specific features.

##### 1) Global Brain Feature Extraction

A Global Anatomical Encoder (built upon Res-UNet [15]) was employed to jointly process T1w, T2w, and FLAIR images, extracting hierarchical structural features. The encoder comprises five downsampling blocks, each structured with two 3 × 3 convolutional layers (with batch normalization and ReLU activation), followed by a 2×2 max pooling layer. This design reduces the spatial resolution from 512×512 to 32×32 while increasing the channel depth from 1 (per modality) to 512, thereby generating a unified global feature map **F**_global_.

##### 2) Multi-Modal Tumor Detection and Encoding

For each modality (T1w, T2w, FLAIR), a dedicated YOLO-based Multi-Modal Tumor detector was employed to localize tumor regions. Each detector was pre-trained on the cerebral tumor datasets, sharing an identical architecture but incorporating modality-specific weights. This enables consistent tumor localization across T1w, T2w, and FLAIR contrasts, addressing modality-specific appearance variations. Each detector’s head output bounding box coordinates and class probabilities, adhering to an Intersection over Union (IoU) threshold of 0.7 for validation. Following detection, tumor regions from all three modalities underwent a standardized processing pipeline: they were first cropped via ROIAlign to a fixed spatial dimension (e.g., 64×64 pixels), then concatenated into a multi-modal tumor patch **P**_tumor_, and finally fed into a shared Tumor Semantic Encoder to generate a tumor-specific feature vector **F**_tumor_. The shared Tumor Semantic Encoder ensures cross-modality feature alignment while reducing parameter redundancy.

##### 3) Feature Fusion and Preliminary Generation

The global brain features **F**_global_ and tumor-specific features **F**_tumor_ were fused via a cross-modal fusion module that employs channel-wise concatenation (**F**_global_ ⊕ **F**_tumor_), followed by a 1×1 convolution to reduce the combined channel dimension to 256. Residual connections were integrated to preserve low-level spatial details and mitigate information loss. The fused features were then upsampled to 512×512 through a Global Anatomical Decoder (five upsampling blocks with skip connections from the Global Anatomical Encoder), producing the preliminary T1C_phase1 prediction.

#### Stage 2: Refinement with Diffusion-Weighted Data

Stage 2 refined tumor boundaries using a hierarchical pipeline initialized with a 4-channel input stack: T1C_phase1, DWI_T1, DWI_T2, and ADC (all 512×512). By propagating features from all three downsampling blocks of the encoder, the hierarchical fusion integrated coarse-to-fine contextual information to enhance boundary precision and tissue contrast efficiency.

##### 1) Cross-Modality Feature Fusion

The Cross-Modality Feature Aggregator processed the 4-channel input stack through sequential operations to generate a fused feature representation. First, a 3×3 convolutional layer expands the channel depth from 4 to 128. A spatial attention mechanism (e.g., spatial dropout with p=0.1) was then utilized to suppress background noise. This pipeline produced a fused feature map **F**_fused_ that encoded both structural (T1C) and diffusive (DWI/ADC) information to be fed into the Local Refinement Encoder.

##### 2) Decoder with Feature Pyramid Network Integration

After the Feature Recovery Unit, data were then processed by the Local Refinement Decoder. This decoder employed three upsampling stages, each paired via skip connections to corresponding downsampling blocks from the Local Refinement Encoder. Feature Pyramid Network (FPN) layers at each stage integrated multi-scale features: low-resolution features (32×32) from the deepest encoder layer captured global tumor morphology, while high-resolution features (256×256) from shallow encoder layers preserved fine-grained details (e.g., microvascular patterns). A final 1×1 convolution layer then mapped features to the T1C intensity values normalized [0, 1].

##### 3) Output Generation

The decoder upscaled features from 32×32 to 512×512 for resolution alignment with stage 1 through bilinear interpolation and residual skip connections, producing the final high-precision T1C_Phase2 image.

### 2.2 Key Architectural Components

The ZeroCEMR framework achieves feature enhancement for multi-modal MRI data and contrast-free T1C image generation through the following core components, focusing on tumor feature separation, inter-stage information retention, and multi-scale feature fusion.

#### 2.2.1 Multi-Modal YOLO Tumor Detection Module

Modality-specific tumor detectors are used to localize lesion regions, leveraging YOLO’s Darknet-53 backbone for multi-scale object detection. Detected tumor regions are cropped via ROIAlign and fed into a shared Tumor Encoder (3-layer 3×3 convolutions) to generate tumor-specific feature vectors. This design avoids dilution of tumor signals by global brain features through a modality-specific detection-shared encoding mechanism, while improving localization robustness using multi-modal contrast consistency.

#### 2.2.3 Tumor Feature Attention Module (SE-Block)

A Squeeze-and-Excitation (SE) block [16] is embedded after the Tumor Semantic Encoder to capture inter-channel dependencies via global average pooling, generating channel attention weights to dynamically adjust the importance of tumor features. This module strengthens tumor-related channels (such as edema signals in FLAIR) and suppresses background noise, improving the model’s sensitivity to small lesions or low-contrast tumors and achieving adaptive calibration of feature spaces.

#### 2.2.3 Multi-Scale FPN UNet

The UNet architecture incorporates an FPN, fusing multi-scale features from corresponding encoder layers through lateral connections at each upsampling stage in the decoder: low-resolution deep features provide tumor semantic information (such as category), while high-resolution shallow features preserve spatial details (such as edges). Combined with pyramid pooling using 3×3 and 5×5 convolutions, the model can handle both 2 mm micro-metastases and 50 mm large-volume tumors, addressing detection biases caused by tumor size heterogeneity.

### 2.3 Training Strategy

ZeroCEMR employws a coarse-to-fine adversarial training strategy [17] across two stages, leveraging a UNetDiscriminatorSN for adversarial learning and a multi-component loss function to balance structural accuracy and perceptual realism.

#### 2.3.1 Stage 1 Training

During Stage 1 training, the input data comprised T1w, T2w, and FLAIR images (512×512) paired with ground-truth T1C images. The objective was to generate a preliminary T1C prediction (O_Stage1_) by optimizing the following loss components.

- **Smooth L1 Loss:** This loss ensured pixel-wise intensity consistency by minimizing the absolute difference between predicted and ground-truth pixel values, facilitating accurate grayscale reproduction. It is defined as:

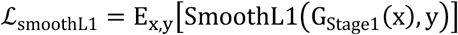
- **SSIM Loss (Structural Similarity Loss):** Designed to preserve structural similarity, this loss quantified the similarity in luminance, contrast, and structural components between the prediction and ground truth, ensuring the integrity of brain anatomical structures. It is defined as:

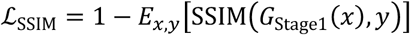
- **Adversarial Loss:** This loss trained the generator to fool the UNetDiscriminatorSN (D) via an adversarial learning framework. By forcing the generator to produce outputs indistinguishable from real T1C images, it enhanced the visual realism and clinical plausibility of the predictions. It is defined as:

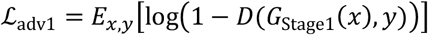

The AdamW optimizer was configured with a weight decay of 0.01 and an initial learning rate of 2e-4, annealed via cosine scheduling to 1e-6 over 100 epochs. This setup balances rapid convergence with fine-grained parameter tuning, leveraging weight regularization to prevent overfitting and cosine annealing to escape local minima, ensuring robust generalization and high-fidelity output generation.

#### 2.3.2 Stage 2 Training

In Stage 2, the input data consisted of the concatenated outputs from Stage 1 (O_Stage1_) combined with DWI and ADC sequences, each paired with corresponding ground-truth T1C images. The primary objective was to refine the preliminary T1C prediction (O_Stage1_) by incorporating additional loss terms.

- **Perceptual Loss:** This loss ensured semantic consistency by aligning high-level features of generated images with those of real images using a pre-trained VGG16 network. It is defined as:

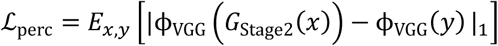
- **Focal Frequency Loss:** This loss enhanced high-frequency detail preservation in the Fourier domain. It is mathematically defined as:

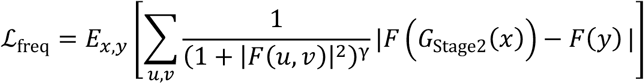

where *F*(·) denotes Fourier transform, (*u, v*) are frequency coordinates, and *γ*= 2 is a focal parameter.
- **Adversarial Loss:** It reinforced realism through the same UNetDiscriminatorSN. The loss is defined as:

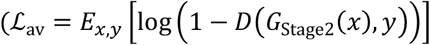
- **Total Loss:** It combines these components into a unified objective:

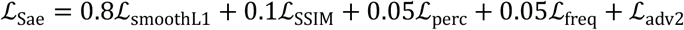

We adopted a optimization strategy by fine-tuning the model on weights obtained from Stage 1 training. Using the AdamW optimizer with a learning rate of 5e-4, we conduct 50 epochs of optimization. This approach leverages pre-learned representations to stabilize training and accelerate convergence, ensuring the model effectively balances semantic correctness, fine-grained detail, and perceptual realism.

#### 2.2.3 Adversarial Training Details

To stabilize the adversarial training process and enforce fine-grained realism in generated outputs, we employed a UNetDiscriminatorSN architecture consisting of seven layers with spectral normalization (SN) applied to all convolutional layers. This discriminator is designed to output a spatial probability map that distinguishes between real and synthetic regions, enabling pixel-level discrimination. Spectral normalization was critical for ensuring Lipschitz continuity in the discriminator’s weight updates, thereby preventing gradient explosion and mode collapse typical in adversarial frameworks.

Data augmentation was systematically applied during both training stages to enhance model generalization and robustness. Specifically, we incorporated random rotations (±15°) to improve orientation invariance, elastic deformations to simulate tissue variability and anatomical flexibility, Gaussian noise injection (σ = [5, 15]) to mitigate sensitivity to imaging artifacts, and Gamma correction (γ = [0.5, 1.5]) to address contrast and illumination inconsistencies. This integrated approach reinforced the generator’s capacity to produce contextually coherent, high-fidelity outputs that adhered to the statistical properties of the training datasets.

### 2.4. Evaluation Metrics

Performance was assessed using PSNR and SSIM to measure the quality of the synthesized T1C images. PSNR measures the quality of a reconstructed image by quantifying the peak signal power relative to the noise power introduced during synthesis. A higher PSNR indicates a better quality of the reconstructed image. The calculation is as below:

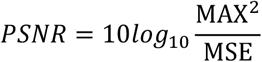

where MAX is defined as the highest possible pixel value within an image, MSE is the average of the squared differences between corresponding pixels of the original and synthetic images.

SSIM assesses the perceptual similarity between two images by considering luminance, contrast, and structural information. With a higher SSIM between the synthesized and ground truth image, the model is better at generating image resembling the reference. It is computed as:

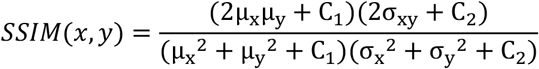

where x and y denote the pre-contrast and post-contrast images respectively, which signifies a high degree of structural consistency, μ is the mean and σ^2^ is the variance and C is the small constant to stabilize the division with weak denominators.

## 3. Result

### 3.1. Datasets

Multi-modal MRI datasets were collected from two sources: the public RSNA-ASNR-MICCAI BraTS 2021 Benchmark (BraTS2021) and a retrospective cohort from a Hong Kong Hospital. The BraTS2021 dataset provides a large, well-curated collection of pre-operative baseline MRI scans from 2,040 patients with gliomas. For this study, we utilized the training cohort, which consists of 1,251 cases. Each case includes co-registered, skull-stripped, and resampled (1 mm^3^ isotropic resolution) volumes of native T1-weighted (T1w), post-contrast T1-weighted (T1C), T2-weighted (T2w), and T2 Fluid Attenuated Inversion Recovery (T2-FLAIR) sequences. Expert neuroradiologists manually annotated and approved the tumor sub-regions, ensuring high-quality ground truth. The dataset’s heterogeneity, stemming from multiple institutions and scanner types, is ideal for training and evaluating the robustness of our model across diverse clinical settings. To complement the BraTS2021 data and provide the additional diffusion-weighted sequences (DWI, ADC) required for our second-stage refinement, we included a multi-modal MRI dataset from Prince of Wales Hospital. This dataset comprises 43 retrospectively enrolled patients with pathologically confirmed glioblastoma who underwent preoperative MRI between 2024-08-21 and 2024-10-28. The inclusion criteria were: (1) adult patients (≥18 years old) with a confirmed diagnosis; (2) availability of a complete preoperative multi-parametric MRI protocol including T1w, T1C, T2w, FLAIR, DWI, and ADC; and (3) MRI examinations meeting diagnostic quality without significant artifacts. Patients with prior surgical intervention, radiotherapy, or chemotherapy were excluded. All images were acquired on 3T Siemens MAGNETOM Prisma scanners. This retrospective study was approved by the Institutional Review Board of Prince of Wales Hospital, which waived the requirement for informed consent. All patient identifiers were removed to ensure complete de-identification before analysis. This dataset was further randomly split into training (30 cases) and testing (13 cases) subsets.

### 3.2. Data Pre-processing

After data collection, MRI images were pre-processed using a standardized pipeline to ensure consistency across modalities and scanners. All sequences (T1w, T2w, FLAIR, DWI, ADC, T1C) were rigidly co-registered to the T1w reference, guaranteeing precise alignment of anatomical structures. Intensity normalization rescaled each slice to [0, 1] by division against its maximum pixel value, preserving intrinsic contrast and standardizing neural network input. Given inconsistencies in the size and resolution across different sequences, all images were resampled to a uniform 512 × 512 pixel dimension using cubic interpolation, which is critical for multi-modal fusion. To augment the training dataset and improve generalization, images underwent rotation (±15°) and horizontal/vertical flipping, simulating minor positioning variations and field-of-view differences while maintaining clinical realism. Noiseless transformations were prioritized to avoid deviating from authentic imaging artifacts. DWI and ADC sequences were corrected for eddy currents and motion artifacts using scanner-specific algorithms, and T1C sequences retained native contrast enhancement patterns post-normalization. Pre-processing was implemented using Python libraries (NumPy, SciPy, MONAI) with GPU acceleration, and a fixed random seed ensured deterministic data splitting and augmentation for reproducibility.

### 3.3. Results

The ZeroCEMR framework achieved exceptional synthesis fidelity, surpassing state-of-the-art methods with a PSNR of 37.82 (95% CI: 37.59–37.99) and SSIM of 0.98 (95% CI: 0.97–0.98). These metrics indicate near-perfect structural similarity and minimal pixel-wise deviation from ground truth and the results approach the theoretical maximum for medical image synthesis tasks. Critically, they demonstrate high-fidelity preservation of both macroscale anatomical features and microscale pathological characteristics essential for lesion detection. As illustrated in Fig. 2, synthetic outputs successfully reconstruct tumor enhancement patterns and delineate precise lesion boundaries. Visual inspection confirms synthetic images are qualitatively indistinguishable from acquired T1C scans in capturing clinically relevant gadolinium uptake dynamics.

**Fig. 2.**
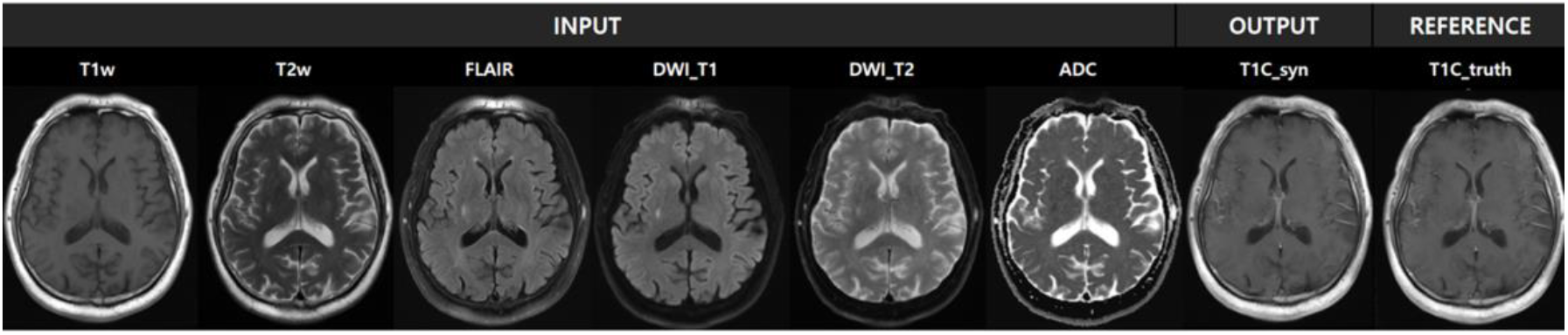
Comparison of the input, output, and reference image

### 3.4 Ablation Studies

As shown in Fig. 3, two steps of ablation yields three architecture utilized in the ablation studies. UnetRe_ serves as the base model; cunetRe builds on unetRe_ by integrating global anatomy encoder, cross-modality feature aggregator, and local refinement encoder for enhanced fusion. And yoloRe, the final framework used in ZeroCEMR, further extends cunetRe by incorporating a multi-modal tumor detector and its corresponding tumor semantic encoder. These three models were trained and tested with the same datasets.

**Fig. 3.**
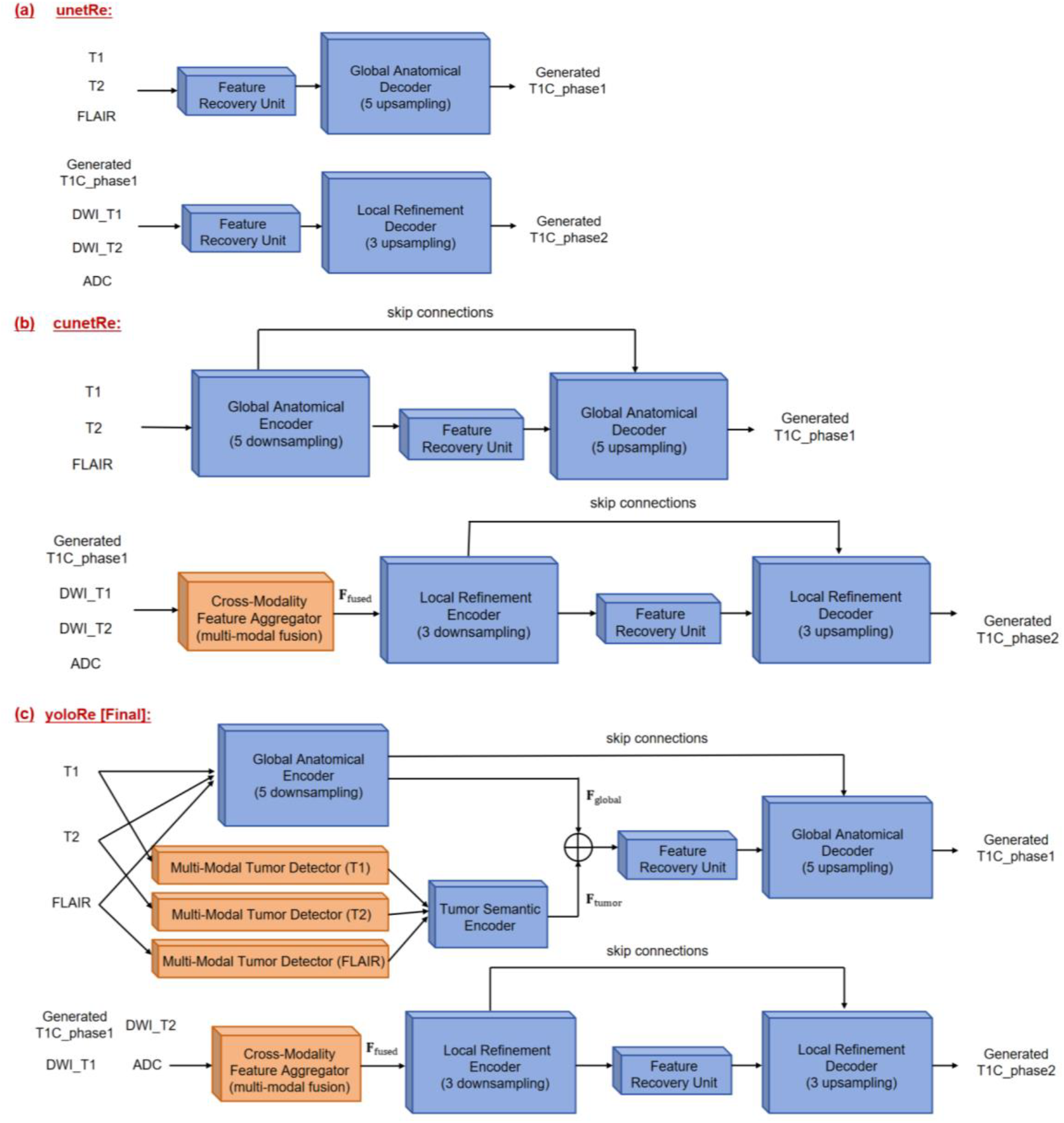
Details of three ablation testing models: (a) unetRe_ is the basic model; (b) cunetRe concatenates global anatomy encoder, cross-modality feature aggregator and local refinement encoder to unetRe_ for fusion; (c) yoloRe subsequently adds multi-modal tumor detector and the corresponding tumor semantic encoder on top of cunetRe.

#### 1) Qualitative result

Three representative cases from the ablation studies are presented in Fig. 4, demonstrating T1C image reconstruction results for tumors varying in size, shape, and anatomical location. Visual comparisons reveal incremental improvements in lesion characterization as model components are sequentially integrated. From unetRe_ to cunetRe, the inclusion of contextual modeling enhances the visibility of fine-scale details in tumor regions, making lesions more distinguishable against surrounding brain tissue. The subsequent transition to the final model yoloRe further refines structural fidelity by integrating multi-modal tumor detector and tumor semantic encoder to avoid tumor feature dilution. This elevates overall image quality to a level comparable with ground truth.

**Fig. 4.**
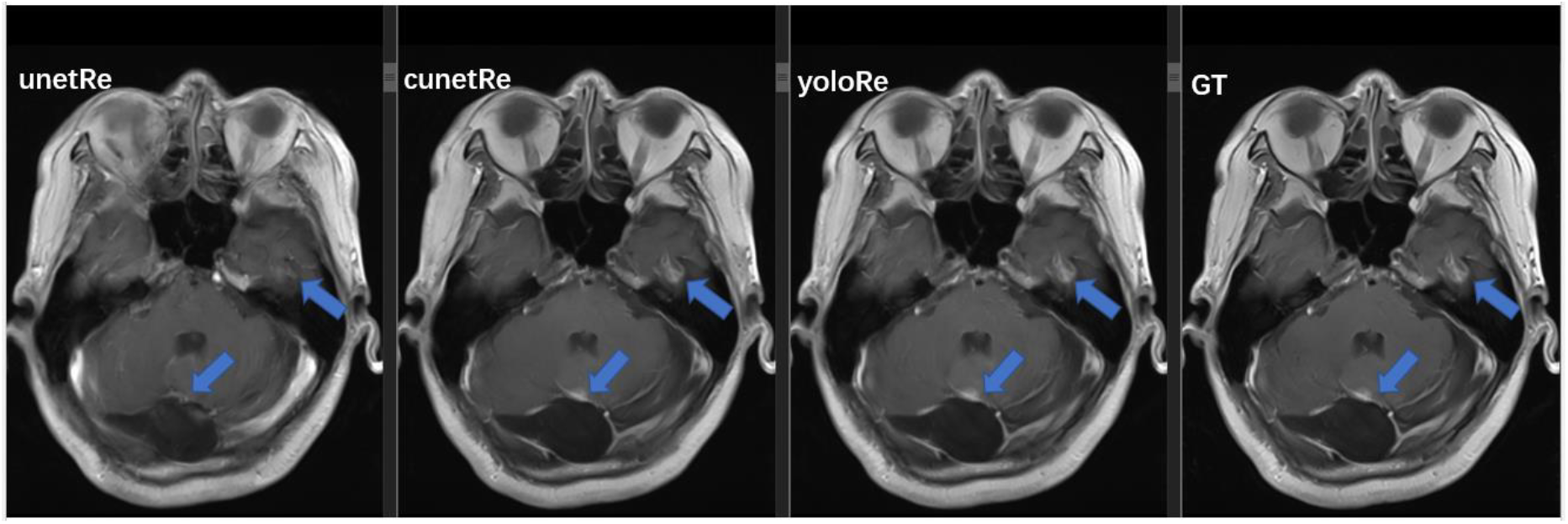
Comparison of three ablation models and the ground truth images. Tumor sensitivity increases with more processing units added, proving the effectiveness of each component in the model.

These results indicate that model accuracy progressively increases with each added component, and the final architecture achieves robust performance across diverse lesion morphologies and brain locations.

#### 2) Quantitative result

Fig. 5 presents the distribution of average PSNR and SSIM values per patient across all reconstruction methods utilized in the experiment. The median PSNR values for unetRe_, cunetRe and yoloRe were 31.66 (95% CI 31.09 - 32.02), 34.64 (95% CI 34.19 - 34.98) and 37.82 (95% CI 37.59 - 37.99). While the median of SSIM for the three methods were 0.94 (95% CI 0.94 - 0.95), 0.96 (95% CI 0.96 - 0.96) and 0.98 (95% CI 0.97 - 0.98) respectively. Both metrics demonstrated progressive improvement across the ablation sequence.

**Fig. 5.**
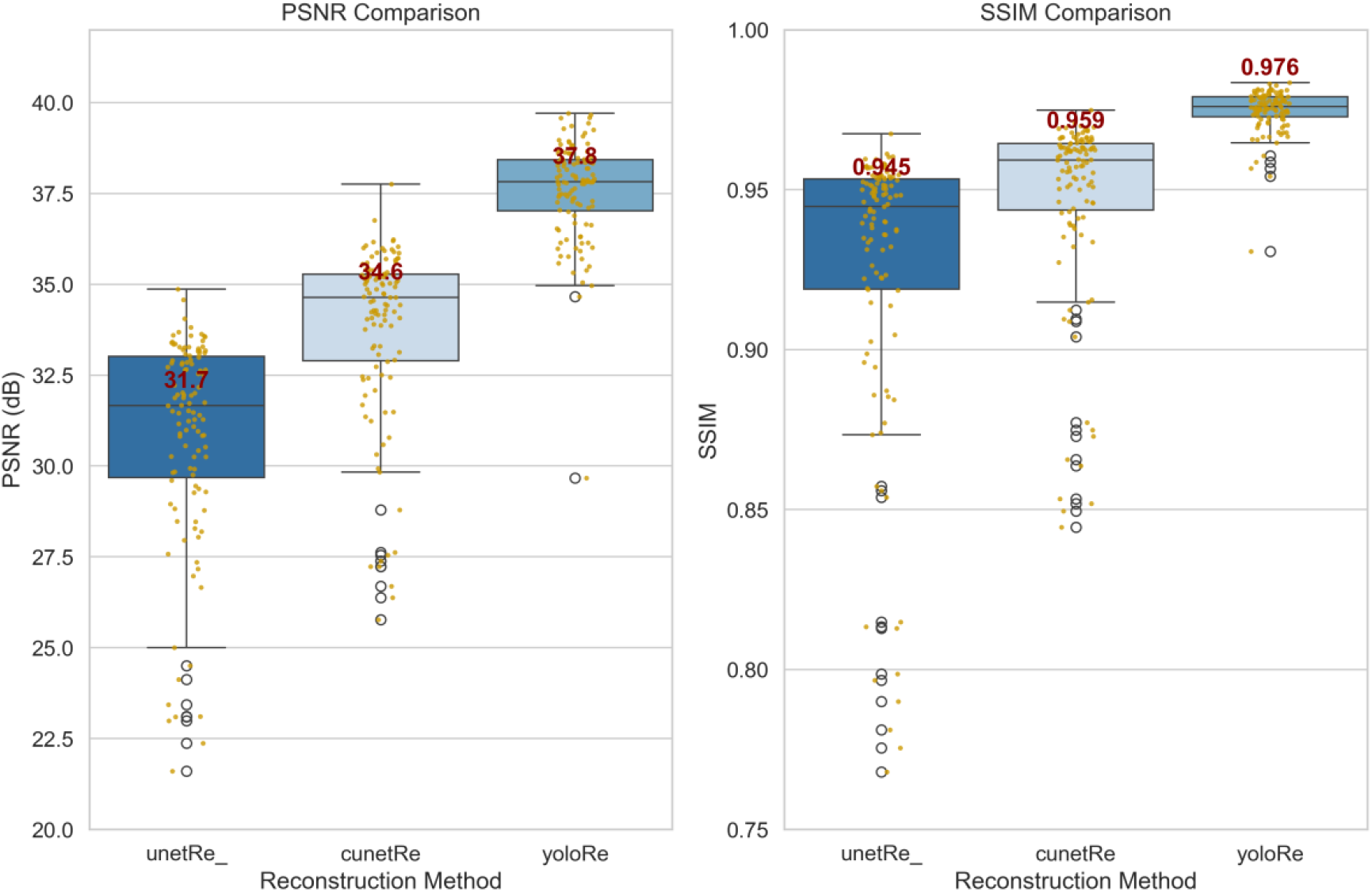
The box and whisker plot for PSNR and SSIM to three reconstruction methods, with the median number labeled in the graph. PSNR = Peak Signal to Noise Ratio, SSIM = Structural Similarity Index.

Statistical analysis confirmed these observations. For PSNR comparison, a Friedman test revealed a global effect (*χ*^*2*^ = 216.05, p < 0.001), prompting post-hoc Dunn’s tests with Bonferroni correction. According to the result of Dunn’s test, the two pairs of methods, there were statistically significant differences between unetRe_/cunetRe (p < 0.001) and cunetRe/yoloRe (p < 0.001). This indicates residual fusion and feature integration significantly enhanced reconstruction performance. For SSIM comparison, parallel analyses yielded consistent results: the Friedman test was significant (p < 0.001), and Dunn’s tests showed all pairwise differences were significant (p < 0.001), with yoloRe outperforming both prior models. The statistical analysis is consistence with the visual assessment of the synthesis image, confirming the incremental performance gains achieved through successive component integration.

Combining both qualitative and quantitative analysis, our ablation studies confirmed the contributions of residual fusion, attention mechanisms, and multi-scale feature integration to the model’s performance. And the studies also suggest that each architectural component contributes uniquely to diagnostic accuracy, and the cumulative improvements culminate in a model capable of generating clinically relevant reconstructions across diverse tumor profiles.

## 4. Discussion

In this work, we propose the multi-modal, two-stage ZeroCEMR framework for synthesizing T1-contrast (T1C) MR images. This approach effectively addresses key challenges in existing methods, particularly low sensitivity in detecting small lesions. Several prior studies have leveraged deep learning for T1C synthesis; we compare our framework with these works in terms of input modalities and model architecture.

Regarding input diversity, Preetha et al. [10] used T1w, T2w, and FLAIR images, while [14] incorporated T1w, T2w, DWI, and ADC data. Zhang et al. [18] trained their model with T1w, T2w, and arterial spin labeling (ASL) data. In contrast, our model accepts five MRI sequences as input. T1-weighted images provide essential anatomical context, enabling the model to learn the normal structure of tissues. T2-weighted images excel at highlighting water-rich abnormalities, such as edema, which are crucial for identifying pathological changes. FLAIR images suppress the signal from cerebrospinal fluid (CSF), enhancing the visibility of lesions adjacent to CSF-filled regions. Critically, we include DWI and ADC map which are rarely utilized in T1C synthesis. They have been shown helpful for cancer diagnosis and tumor response assessment [19] due to their capability to demonstrate water molecule diffusion and reveal microstructural changes in lesion tissues. By combining these diverse modalities, our model gains a holistic understanding of tissues, allowing it to generate post-contrast T1w images that accurately depict both normal anatomical features and disease-related alterations.

Previous attempts at synthesizing post-contrast T1w images from pre-contrast ones have faced significant limitations, primarily manifested as blurred pathology representation and low sensitivity to small lesions [20–22]. For instance, a previous model purely based on Pix2pixHD GAN attained an SSIM of 0.86 (95% CI 0.85 - 0.87) and a PSNR of 26.35 (95% CI 25.94 - 26.76) [18]. Another work built on a simple 3D dCNN reported an SSIM of 0.82 [11]. A comparison of 3D CNN and conditional GAN (cGAN) in [10] yielded SSIMs of 0.809 (95% CI: 0.808–0.811) and 0.827 (95% CI: 0.826 – 0.829), respectively. To overcome these limitations, our ZeroCEMR framework capitalizes on the strengths of a Feature Pyramid Network integrated with Unet, effectively fusing multi-scale features to capture both semantic and spatial information. Additionally, adversarial training enforces fine-grained realism in the generated images. Our key innovative Multi-Modal YOLO Tumor Detection Module prevents signal dilution and boosts lesion localization accuracy. As a result, our model demonstrates a remarkable improvement, achieving an SSIM of 0.98 (95% CI 0.97 - 0.98) and a PSNR of 37.82 (95% CI 37.59 - 37.99), significantly outperforming previous approaches.

While the ZeroCEMR framework demonstrates promising performance, several limitations warrant attention for future advancement. First, our training data for phrase 2 was acquired primarily from scanners at solely one medical center, limiting diversity in device protocols and patient populations. Variations in MRI hardware (e.g., field strength, coil design) and acquisition protocols across institutions can introduce technical heterogeneity, potentially affecting the model’s cross-device robustness. Expanding the dataset to include multi-center, multi-vendor data would enhance generalizability and reduce bias toward specific imaging environments. Second, the framework currently operates on 2D image slices, overlooking the spatial continuity and inter-slice relationships inherent in 3D volumetric data. Integrating 3D convolutional architectures or transformer-based models capable of leveraging inter-slice contextual information could improve the accuracy of tumor boundary delineation and small lesion visualization [12], particularly in cases where lesions span multiple slices. Third, the training data is primarily focused on a subset of common brain diseases, which may limit the model’s ability to generalize to rare or atypical lesions with unique morphological or diffusion characteristics. Addressing this requires incorporating diverse pathological subtypes into the dataset and exploring techniques like transfer learning or few-shot learning to enhance adaptability to underrepresented conditions.

Looking ahead, the framework’s utility extends beyond diagnostic imaging to longitudinal clinical applications. By generating high-fidelity post-contrast T1C images from pre-contrast multi-modal data, the model could facilitate disease progression monitoring by enabling consistent, standardized comparisons of tumor morphology over time without the need to inject GBCA each time. This significantly reduces the toxicity accumulation. In addition, integrating the framework with dynamic contrast-enhanced (DCE)-MRI analysis or physiological models could support treatment response assessment, such as predicting patient outcomes following chemotherapy or radiation therapy. Future work might also explore combining generated T1C images with other advanced modalities (e.g., positron emission tomography [PET]) to create multi-modal synthetic datasets for training downstream tasks like image segmentation or survival prediction. Ultimately, advancing toward fully automated, clinically deployable systems will require addressing scalability, real-time inference speed, and regulatory compliance. Collaborations with multi-institutional consortia to curate diverse datasets and validate performance across global populations will be pivotal in translating this research into impactful clinical tools.

## 5. Conclusion

In conclusion, the ZeroCEMR framework establishes a new state-of-the-art for synthesizing T1C MRI by integrating multi-parametric inputs with a lesion-aware, two-stage architecture. Our method significantly outperforms existing approaches, achieving superior quantitative metrics and high-fidelity lesion delineation. This work provides a robust and clinically promising pathway toward gadolinium-free quantitative neuro-oncology, effectively bridging a critical gap between advanced deep learning innovation and practical clinical MRI applications.

## Data Availability

All data produced in the present study are available upon reasonable request to the authors

## References

[1] Bauer, S., Wiest, R., Nolte, L. P., & Reyes, M. (2013). A survey of MRI-based medical image analysis for brain tumor studies. Physics in Medicine & Biology, 58(13), R97.

[2] Choi, J. W., & Moon, W. J. (2019). Gadolinium deposition in the brain: current updates. Korean journal of radiology, 20(1), 134–147.

[3] Fraum, T. J., Ludwig, D. R., Bashir, M. R., & Fowler, K. J. (2017). Gadolinium-based contrast agents: a comprehensive risk assessment. Journal of Magnetic Resonance Imaging, 46(2), 338–353.

[4] Runge, V. M. (2017). Critical questions regarding gadolinium deposition in the brain and body after injections of the gadolinium-based contrast agents, safety, and clinical recommendations in consideration of the EMA’s pharmacovigilance and risk assessment committee recommendation for suspension of the marketing authorizations for 4 linear agents. Investigative radiology, 52(6), 317–323.

[5] Gulani, V., Calamante, F., Shellock, F. G., Kanal, E., & Reeder, S. B. (2017). Gadolinium deposition in the brain: summary of evidence and recommendations. The Lancet Neurology, 16(7), 564–570.

[6] European Medicines Agency. (2017). EMA’s final opinion confirms restrictions on use of linear gadolinium agents in body scans. EMA website.

[7] Gong, E., Pauly, J. M., Wintermark, M., & Zaharchuk, G. (2018). Deep learning enables reduced gadolinium dose for contrast-enhanced brain MRI. Journal of magnetic resonance imaging, 48(2), 330–340.

[8] Müller-Franzes, G., Huck, L., Tayebi Arasteh, S., Khader, F., Han, T., Schulz, V., … & Truhn, D. (2023). Using machine learning to reduce the need for contrast agents in breast MRI through synthetic images. Radiology, 307(3), e222211.

[9] Porter, K. K., King, A., Galgano, S. J., Sherrer, R. L., Gordetsky, J. B., & Rais-Bahrami, S. (2020). Financial implications of biparametric prostate MRI. Prostate cancer and prostatic diseases, 23(1), 88–93.

[10] Preetha, C. J., Meredig, H., Brugnara, G., Mahmutoglu, M. A., Foltyn, M., Isensee, F., … & Vollmuth, P. (2021). Deep-learning-based synthesis of post-contrast T1-weighted MRI for tumour response assessment in neuro-oncology: a multicentre, retrospective cohort study. The Lancet Digital Health, 3(12), e784–e794.

[11] Calabrese, E., Rudie, J. D., Rauschecker, A. M., Villanueva-Meyer, J. E., & Cha, S. (2021). Feasibility of simulated postcontrast MRI of glioblastomas and lower-grade gliomas by using three-dimensional fully convolutional neural networks. Radiology: Artificial Intelligence, 3(5), e200276.

[12] Chen, C., Raymond, C., Speier, W., Jin, X., Cloughesy, T. F., Enzmann, D., … & Arnold, C. W. (2022). Synthesizing MR image contrast enhancement using 3D high-resolution ConvNets. IEEE Transactions on Biomedical Engineering, 70(2), 401–412.

[13] Synthetic Post-Contrast Imaging through Artificial Intelligence: Clinical Applications of Virtual and Augmented Contrast Media.

[14] Huang, H., Mo, J., Ding, Z., Peng, X., Liu, R., Zhuang, D., … & Qiu, Y. (2025). Deep Learning to Simulate Contrast-Enhanced MRI for Evaluating Suspected Prostate Cancer. Radiology, 314(1), e240238.

[15] Ronneberger, O., Fischer, P., & Brox, T. (2015, October). U-net: Convolutional networks for biomedical image segmentation. In International Conference on Medical image computing and computer-assisted intervention (pp. 234–241). Cham: Springer international publishing.

[16] Vaswani, A., Shazeer, N., Parmar, N., Uszkoreit, J., Jones, L., Gomez, A. N., … & Polosukhin, I. (2017). Attention is all you need. Advances in neural information processing systems, 30.

[17] Goodfellow, I. J., Pouget-Abadie, J., Mirza, M., Xu, B., Warde-Farley, D., Ozair, S., … & Bengio, Y. (2014). Generative adversarial nets. Advances in neural information processing systems, 27

[18] Zhang, Y., Huang, Y., Xiong, X., Liu, Y., & Qi, J. (2024). A multi-task generative model for simultaneous post-contrast MR image synthesis and brainstem glioma segmentation. Magnetic Resonance Imaging, 113, 110210.

[19] Afaq, A., Andreou, A., & Koh, D. M. (2010). Diffusion-weighted magnetic resonance imaging for tumour response assessment: why, when and how?. Cancer Imaging, 10(1A), S179.

[20] Osuala, R., Joshi, S., Tsirikoglou, A., Garrucho, L., Pinaya, W. H., Diaz, O., & Lekadir, K. (2024, April). Pre-to post-contrast breast MRI synthesis for enhanced tumour segmentation. In Medical Imaging 2024: Image Processing (Vol. 12926, pp. 226–237). SPIE.

[21] Tabassum, M., Rana, P., Suero Molina, E., Di Ieva, A., & Liu, S. (2024, July). Cross-Modality Synthesis of T1c MRI from Non-contrast Images Using GANs: Implications for Brain Tumor Research. In International Conference on Artificial Intelligence in Medicine (pp. 60–69). Cham: Springer Nature Switzerland.

[22] Zhou, Z., Arefan, D., Zuley, M. L., Sumkin, J. H., & Wu, S. (2024, April). Quantifying the quality of GAN-synthesized images: a study on synthesizing post-contrast sequences from precontrast sequences in breast DCE-MRI. In Medical Imaging 2024: Computer-Aided Diagnosis (Vol. 12927, pp. 546–550). SPIE.

